# Prioritizing context-specific genetic risk mechanisms in 11 solid cancers

**DOI:** 10.1101/2025.10.30.25339145

**Authors:** Xueyao Wu, Artem Kim, Charles E Breeze, Tracy A O’Mara, Dhanya Ramachandran, Thilo Dörk, Stella Koutros, Nathaniel Rothman, Ludmila Prokunina-Olsson, Nicholas Mancuso, Sara Lindström, Peter Kraft

## Abstract

**Background:** While genome-wide association studies (GWAS) have identified hundreds of cancer-associated genetic variants, the specific biological contexts where these variants exert their effects remain largely unknown. We aimed to prioritize context-specific genetic risk mechanisms for 11 solid cancers at both genome-wide and single-variant resolutions.

**Methods:** We integrated cancer GWAS summary statistics from European ancestry samples (avg. n cases=47,856) with ∼1,500 context-specific annotations representing candidate *cis*-regulatory elements. For genome-wide analysis, we applied CT-FM, a method that leverages heritability enrichment estimates and an annotation correlation matrix to select likely disease-relevant biological contexts. After identifying putative causal SNPs (PIP≥0.5) via functionally informed fine-mapping, we used CT-FM-SNP to identify relevant contexts for individual variants. A combined SNP-to-gene framework was applied to construct putative {regulatory SNP-context-gene-cancer} quadruplets.

**Results:** Stratified LD score regression analysis identified 52 annotations with significant heritability enrichment (Bonferroni-corrected *P*≤0.05). CT-FM prioritized four high-confidence (PIP≥0.5) biological contexts: mammary luminal epithelial cells for breast cancer, a prostate cancer epithelial cell line (VCaP) for prostate cancer, and bulk tumor tissue contexts for colorectal and renal cancers. Variant-level analysis of hundreds of putatively causal SNPs corroborated these findings and identified additional high-confidence contexts for other malignancies, including estrogen receptor-negative breast cancer and bladder cancer. A total of 489 putative regulatory quadruplets were constructed, proposing specific molecular mechanisms underlying the observed GWAS signals.

**Conclusion:** These findings advance our understanding of genetic susceptibility to different cancers. Future work in larger, more diverse GWAS, coupled with more comprehensive annotation atlases, is essential to expand upon and validate our results.

## Introduction

Cancer remains a leading cause of morbidity and mortality worldwide, with a global burden that is expected to rise significantly in the coming decades. This highlights the urgent need for improved prevention, early detection, and treatment strategies.^1,2^ While genome-wide association studies (GWAS) have identified hundreds of genetic variants associated with cancer susceptibility, the vast majority of these variants are located in non-coding regions of the genome.^3^ Translating these associations into biological function and clinical relevance presents a formidable challenge, central to which is pinpointing the causal variants and understanding their context-specific regulatory mechanisms.^4^

Cancer is driven by complex interactions among diverse cell populations.^5^ Genetic risk variants likely exert their effects by perturbing gene regulatory networks within specific contexts,^6,7^ ranging from individual cell types to complex tissue environments comprising multiple cell populations, such as the tumor microenvironment (TME).^8^ For instance, breast cancer risk variant rs2981578 near the *FGFR2* gene selectively alters the binding of FOXA1, a pioneer factor critical for estrogen receptor (ER) function expressed in only a subset of luminal epithelial cells within the human mammary gland.^9^ Analysis of thousands of lung cancer variants revealed that a majority of the risk variants found in regulatory regions were specific to a single cell-type category, with epithelial and immune cells emerging as the most significant contributors to lung cancer susceptibility.^10^ Understanding the biological contexts where cancer-related genetic associations individually and collectively alter gene regulation is therefore critical for unraveling the cellular origins of cancer, prioritizing experimental validation studies, and informing the development of targeted therapies.^11^

Large-scale functional genomics initiatives, including the ENCyclopedia of DNA Elements (ENCODE4) and The Cancer Genome Atlas (TCGA) projects,^12,13^ have generated comprehensive epigenomic profiles across diverse tissue and cellular environments, creating opportunities for integration with GWAS data to elucidate context-specific mechanisms in cancer genetics. Statistical methods such as Stratified Linkage Disequilibrium Score Regression (S-LDSC)^14,15^ leverage these functional annotations for partitioning heritability and identifying disease-relevant biological contexts. However, when evaluating each annotation independently, S-LDSC cannot distinguish truly causal from ‘tagging’ associations arising from context-shared regulatory patterns (e.g., different immune cell types or epithelial cells from different tissues).^16,17^ Consequently, it frequently identifies multiple, highly correlated annotations as statistically significant, substantially obscuring the primary drivers of heritability signals.^18^ On the other hand, performing S-LDSC while mutually adjusting for a large number of context-specific annotations is computationally intractable. This challenge underscores the need for computational frameworks that explicitly model shared regulatory architectures across diverse contexts to enable more precise identification of the biological processes driving cancer development.^19,20^

To address these limitations, we implemented an integrative statistical fine-mapping framework to systematically prioritize the biological contexts underlying genetic risk for 11 solid cancers. Our approach centers on Cell-Type Fine-Mapping (CT-FM) and its single-variant extension, CT-FM-SNP, which adapt established SNP fine-mapping principles to the prioritization of cell-type and tissue contexts by jointly modeling heritability enrichments across annotations.^20^ While these methods have demonstrated success in prioritizing cellular contexts and disentangling correlated signals for multiple continuous traits, their application to cancer and other complex diseases remains limited. To further enhance precision, we employed an advanced, functionally informed SNP fine-mapping strategy to generate refined sets of putative causal variants for each cancer type, which serve as high-confidence input for CT-FM-SNP analysis.^21^ By linking these context-specific variants to their putative target genes, we constructed end-to-end hypotheses that specify the likely causal SNP, its functional context, the gene it regulates, and the associated cancer. Through the integration of these complementary approaches, we sought to dissect the genetic architecture of cancer susceptibility, identify putative regulatory mechanisms, and establish a foundation for future experimental validation and therapeutic target discovery. The overall study design is presented in **Figure 1**.

**Figure 1.**
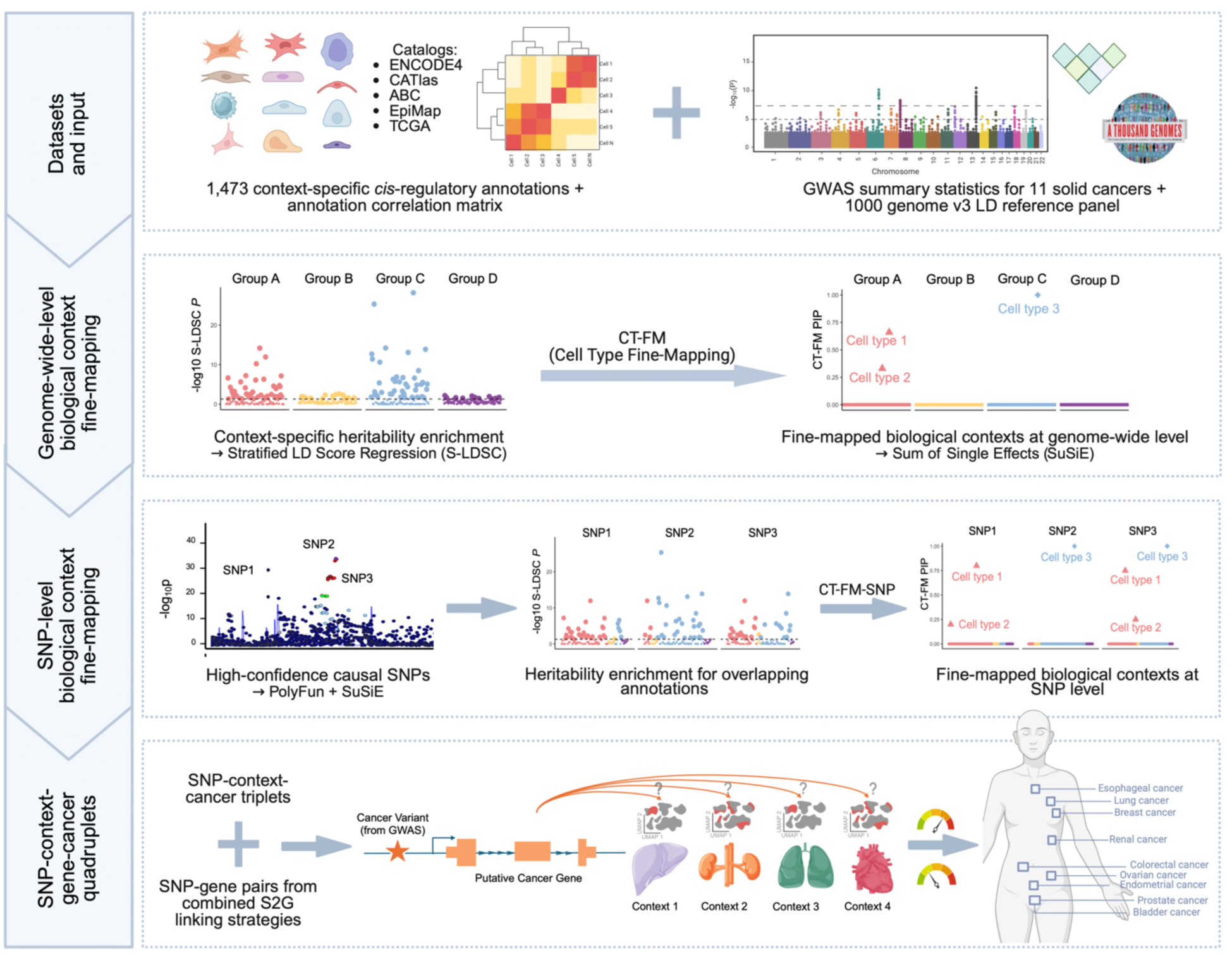
Overall study design. This schematic illustrates the integrative statistical fine-mapping framework employed to prioritize context-specific genetic risk mechanisms in 11 solid cancers. The workflow begins with the integration of large-scale GWAS summary statistics and an atlas of ∼1,500 context-specific *cis*-regulatory annotations. Cell-Type Fine-Mapping (CT-FM) is applied for genome-wide prioritization of biological contexts. Concurrently, functionally informed SNP fine-mapping identifies high-confidence causal variants. These variants then serve as input for CT-FM-SNP to pinpoint relevant contexts at a single-variant resolution. Finally, a combined SNP-to-gene (cS2G) framework links context-specific variants to their putative target genes, culminating in the construction of {regulatory SNP-context-gene-cancer} quadruplets.

## Materials and Methods

### Context-specific SNP-annotations

Our dataset includes 1,473 SNP annotations, each defining a candidate *cis*-regulatory element in a specific biological context (**Table S1**). This collection was assembled by first incorporating 927 cell-type-specific annotations previously curated for the original CT-FM framework from the fourth phase of ENCODE (ENCODE4),^22^ Cis-element Atlas (CATlas),^23^ and Activity-by-Contact (ABC) model.^24^ We then expanded this library with 523 additional cell-type-specific annotations, including cell lines and cancer-related cell types from the original consortia to enhance the scope for cancer-specific analysis, as well as annotations from Epigenome Integration across Multiple Annotation Projects (Epimap)^25^ to broaden overall cellular diversity. We also integrated 23 bulk tumor annotations from TCGA.^13^ Although cell-type-specific data from primary tumors are not widely available, these bulk tumor profiles are essential as they capture the aggregate regulatory landscape of the *in vivo* tumor ecosystem. Our final library therefore combines the cellular specificity of primary cells and cell lines (n = 1,450) with the disease relevance of primary tumor tissues (n = 23). Detailed information on annotation data types, curation and quality control procedures is available in the **Supplementary Methods**.

### GWAS summary statistics

We assembled GWAS summary statistics from European ancestry samples for 11 cancer types. The datasets encompassed breast cancer (total n=228,951; 122,977 cases), with subtype-specific data for estrogen receptor-positive (ER+; 69,501 cases) and estrogen receptor-negative (ER–; 21,468 cases) breast cancer; endometrial cancer (n= 244,644; 16,078 cases); ovarian cancer (n=63,347; 22,406 cases); prostate cancer (n=726,828; 122,188 cases); colorectal cancer (n=185,886; 78,743 cases); esophageal cancer (n=21,271; 4,112 cases); lung cancer (n=85,716; 29,266 cases); renal cancer (n=769,475; 25,890 cases); and bladder cancer (n=357,292; 13,790 cases). Most cancer GWAS datasets were imputed using 1000 Genomes Project (1KGP) reference panels. Study characteristics and participant information are summarized in **Table 1** with further details available in the original cancer-specific publications.^26–34^

**Table 1.** Overview of cancer GWAS datasets.

| **Cancer type** | **Reference (PMID)** | **Release year** | **Cases** | **Controls** | **Ancestry** | **SNPs after QC** | **Imputation reference panel** |
| --- | --- | --- | --- | --- | --- | --- | --- |
| Breast |  |  | 122,977 |  |  | 7,370,487 |  |
| ER+ breast | 29059683 | 2017 | 69,501 | 105,974 | European | 7,370,392 | 1000G Phase 3 |
| ER– breast |  |  | 21,468 |  |  | 7,368,117 |  |
| Endometrial | 40633141 | 2025 | 16,078 | 228,566 | European | 7,254,547 | 1000G Phase 3, UK10K, Estonian and Finnish population-specific imputation references |
| Ovarian (epithelial) | 28346442 | 2017 | 22,406 | 40,941 | European | 7,260,260 | 1000G Phase 3 |
| Prostate | 37945903 | 2023 | 122,188 | 604,640 | European | 8,040,608 | 1000G phase 3, TOPMed freeze 5, HRC, UK10K, or SISu v3 |
| Colorectal | 36539618 | 2022 | 78,743 | 107,143 | European | 7,346,098 | 1000G Phase 1/ Phase 3, HRC, UK10K, or SISu |
| Esophageal | 27527254 | 2016 | 4,112 | 17,159 | European | 7,572,848 | 1000G Phase 1 |
| Lung | 28604730 | 2017 | 29,266 | 56,450 | European | 6,314,046 | 1000G Phase 1/ Phase 3 |
| Renal | 38671320 | 2024 | 25,890 | 743,585 | European | 8,000,795 | TOPMed imputation version R2, HRC, UK10K |
| Bladder | 37210288 | 2023 | 13,790 | 343,502 | European | 6,750,997 | HRC release 1.1, 1000G Phase 3 |

We applied the following quality control filters to each GWAS dataset. We removed variants on sex chromosomes, multi-allelic SNPs, and ambiguous variants; variants with low imputation quality (INFO score < 0.9), low minor allele frequency [MAF] < 0.005, extreme effect sizes (|BETA| > 3), or invalid *P*-values (*P*-value ≤ 0 or > 1); and variants sharing identical physical positions. We harmonized all datasets to the hg19 genome build. Finally, we aligned the datasets with the 1KGP v3 European reference panel, excluding variants absent from the panel, variants with MAF < 0.005 in the reference panel, and variants exhibiting significant allele frequency discrepancies (MAF difference > 0.2) between the GWAS and the reference panel population.

### CT-FM analysis

We utilized CT-FM to prioritize biological contexts underlying cancer-specific GWAS signals.^20^ CT-FM first applies S-LDSC, using LD scores from the 1KGP v3 European reference panel and adjusting for baseline genomic features, to estimate marginal effects of each context-specific annotation on SNP heritability and generate a genome-wide heritability enrichment Z-score for each cancer-context pair.^15^ We used 1/(1/Ncase + 1/Ncontrol) as the effective sample size in the S-LDSC analysis (see rationale in the supplementary note of ^35^). Using enrichment estimates with positive z-scores and a correlation matrix of these annotations (i.e., correlations between the baseline-adjusted LD scores of context-specific annotations)^20^, CT-FM then employs the Sum of Single Effects (SuSiE) model to fine-map annotations, assuming a maximum of 10 independent causal contexts (L = 10, default setting of CT-FM).^36,37^ This process generates (i) annotation-specific posterior inclusion probabilities (PIPs), reflecting the probability of an annotation being a primary driver of heritability enrichment, and (ii) credible sets, where each set represents an independent signal consisting of the smallest group of correlated annotations (with a minimum absolute pairwise correlation ≥ 0.192, the average correlation of the annotation matrix) that together have a 95% probability of including the true causal context. We identified annotations showing statistically significant enrichment in S-LDSC using both Bonferroni-corrected and FDR-corrected *P*-value thresholds of 0.05 within each cancer. Following the original CT-FM framework, we defined high-confidence biological contexts as those with PIPs ≥ 0.5. To increase our sensitivity to identify candidate biological contexts, we also used a more lenient threshold of 0.05 ≤ PIPs < 0.5.

### SNP-fine-mapping analysis

We conducted functionally informed SNP fine-mapping using PolyFun integrated with SuSiE to identify high-confidence causal variants for each cancer.^21^ PolyFun leverages genome-wide functional annotations including coding, conserved, regulatory, and LD-related genomic features to compute prior causal probabilities for variants, proportional to their predicted per-SNP heritability contributions. These annotation-derived priors were then incorporated into the SuSiE fine-mapping algorithm to improve causal variant identification. For our analysis, we utilized a set of pre-computed PolyFun prior probabilities derived from a meta-analysis of 15 genetically uncorrelated UK Biobank traits, covering approximately 19 million imputed SNPs with MAF > 0.1%. Using these functional priors, we applied SuSiE fine-mapping to overlapping 3MB regions across the genome, restricting our analysis to those containing at least one genome-wide significant (P < 5×10^−8^) SNP and assuming 5 causal SNPs in each region (L = 5, default setting of PolyFun). To resolve duplicate analyses for SNPs present in multiple regions, we retained only the result from the region where the SNP was most centrally located. The 1KGP European reference panel was used for LD information. Effective sample size was calculated as Neff = 4/(1/Ncase + 1/Ncontrol), following the approach adopted by the original publication.^21^ High-confidence causal variants were defined with a PIP threshold of ≥ 0.50. The resulting set of variants was then used as input for CT-FM-SNP.

### CT-FM-SNP analysis

To link specific candidate variants to their most relevant biological contexts, we further implemented CT-FM-SNP for each cancer.^20^ CT-FM-SNP differs from CT-FM by restricting the SuSiE fine-mapping analysis to subsets of context-specific annotations that overlap with a given variant. We used the cancer-specific high-confidence causal variants identified through PolyFun+SuSiE fine-mapping as inputs. CT-FM-SNP first identified the subset of annotations that overlapped with the variant and then extracted the genome-wide heritability enrichment estimates for these annotations, as previously calculated by CT-FM. For variants overlapping at least two annotations with positive enrichment z-scores, SuSiE fine-mapping was applied using these variant-restricted annotations and the corresponding annotation correlation matrix, outputting variant-specific PIPs and credible sets for each annotation-cancer pair. As previously, high-confidence biological contexts were defined as those with PIPs ≥ 0.5. We did not focus on candidate contexts at the single-variant level given that the model was restricted to annotations with direct SNP overlap, a scenario where a lenient threshold might elevate the risk of false-positive findings.

### Identification of {regulatory SNP-context-gene-cancer} quadruplets

To generate specific testable hypotheses, we integrated our SNP-level context results with a comprehensive SNP-to-gene (S2G) mapping framework to identify {regulatory SNP-context-gene-cancer} quadruplets. Our approach employed the combined S2G (cS2G) framework which integrates multiple S2G linking strategies to identify putative target genes for regulatory SNPs.^38^ The cS2G framework incorporates 10 functionally informed S2G strategies, such as exon-based, promoter-based, fine-mapped *cis*-eQTLs, enhancer-gene linking, and chromatin interaction data, to assign linking scores to SNP-gene pairs. For our analysis, we utilized pre-computed cS2G linking scores derived from the 1KGP European reference panel. To form putative quadruplets, we systematically linked the SNP-context pairs that passed our high-confidence threshold (CT-FM-SNP PIP ≥ 0.5) with high-confidence (cS2G score ≥ 0.5) SNP-gene pairs from the cS2G framework.

### Sensitivity analyses excluding bulk tumor annotations

To assess the robustness of our findings in the absence of bulk tumor annotations, we performed sensitivity analyses by repeating all analyses (CT-FM, CT-FM-SNP, and quadruplet construction) after excluding the 23 TCGA bulk tumor annotations.

## Results

### Identification of context-specific heritability enrichment

In total, S-LDSC identified 52 biological contexts showing significant heritability enrichment (Bonferroni-corrected *P* ≤ 0.05) across five cancer types: overall breast cancer, ER+ breast cancer, prostate cancer, colorectal cancer, and renal cancer. At a less stringent significance threshold (q-value ≤ 0.05), S-LDSC identified 19, 13, 82, 22, and 5 contexts for overall breast, ER+ breast, prostate, colorectal, and renal cancers, respectively, totaling 141 contexts (**Figure 2**).

**Figure 2.**
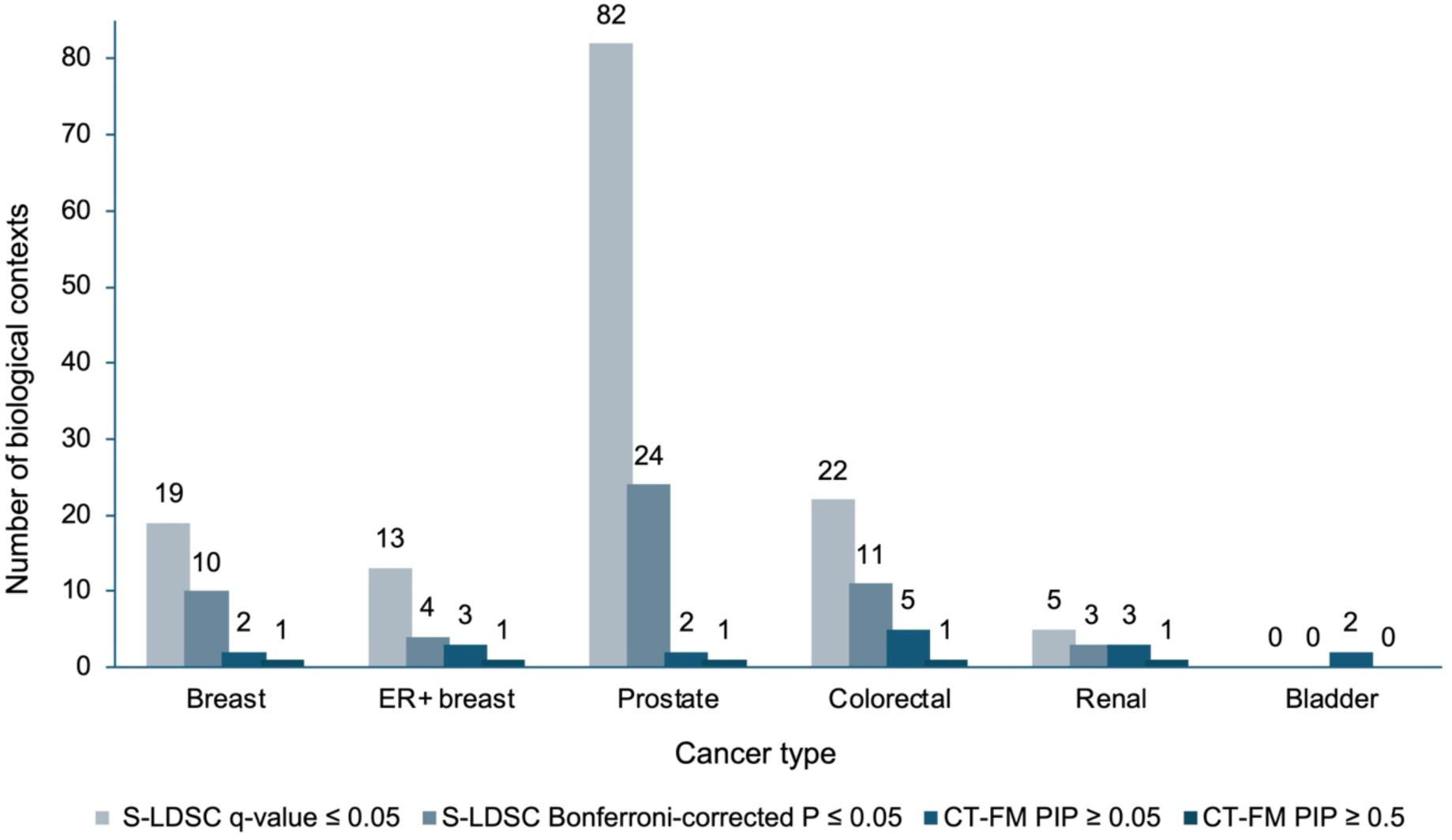
Statistical prioritization of biological contexts across six solid cancer types. Bar chart showing the number of annotations identified for six cancer types (breast, ER+ breast, prostate, colorectal, renal, and bladder) that demonstrated significant genome-wide enrichment signals. Annotations are categorized by increasing levels of statistical stringency: from annotations with stratified linkage disequilibrium score regression (S-LDSC) false discovery rate (FDR) q-value ≤ 0.05 to those meeting Bonferroni-corrected significance (*P* ≤ 0.05) in S-LDSC analysis; and from candidate biological contexts with Cell-Type Fine-Mapping (CT-FM) posterior inclusion probability (PIP) ≥ 0.05 to high-confidence contexts with CT-FM PIP ≥ 0.5.

### Fine-mapping biological contexts at genome-wide resolution

CT-FM substantially refined the initial S-LDSC results (**Table 2**, **Figure 2**), identifying four contexts achieved high confidence (PIPs ≥ 0.5): mammary luminal epithelial cells for both overall and ER+ breast cancer (PIP = 0.62-0.75), VCaP prostate adenocarcinoma epithelial cell line for prostate cancer (PIP = 1.00), colon adenocarcinoma (COAD) bulk tissue for colorectal cancer (PIP = 1.00), and kidney renal clear cell carcinoma (KIRC) bulk tissue for renal cancer (PIP = 0.73).

**Table 2.** High-confidence (PIP ≥ 0.5) and candidate (PIP ≥ 0.05) biological contexts identified by genome-wide context fine-mapping.

| Cancer type | Biological context | Functional category | S-LDSC Z-score | CT-FM PIP | Credible set size |
| --- | --- | --- | --- | --- | --- |
| Breast | **Mammary Luminal Epithelial Cell** | Primary Epithelial | 5.30 | 0.62 | 3 |
|  | Mammary Basal Epithelial Cell | Primary Epithelial | 5.16 | 0.32 | 3 |
| ER+ breast | **Mammary Luminal Epithelial Cell** | Primary Epithelial | 5.15 | 0.75 | 3 |
|  | Mammary Luminal Adenocarcinoma Epithelial Cell (T47D) | Cancer Cell Line | 4.77 | 0.12 | 3 |
|  | Mammary Basal Epithelial Cell | Primary Epithelial | 4.74 | 0.11 | 3 |
| Prostate | **Prostate Adenocarcinoma Epithelial Cell (VCaP)** | Cancer Cell Line | 7.84 | 1.00 | 1 |
|  | Prostate adenocarcinoma (PRAD) | Bulk Tumor Tissue | 7.19 | 0.35 | NA |
| Colorectal | **Colon Adenocarcinoma (COAD)** | Bulk Tumor Tissue | 6.45 | 1.00 | 1 |
|  | Connective Tissue Mesenchymal Stem Cell | Mesenchymal/Stromal | 4.34 | 0.17 | NA |
|  | Lung Fibroblast (WI-38) | Fibroblast Cell Line | 4.29 | 0.14 | NA |
|  | Penis Fibroblast Myc-Immortalized | Immortalized Stromal | 4.14 | 0.07 | NA |
|  | Placenta Villous Mesenchymal Fibroblast | Mesenchymal/Stromal | 4.00 | 0.05 | NA |
| Renal | **Kidney Renal Clear Cell Carcinoma (KIRC)** | Bulk Tumor Tissue | 4.69 | 0.73 | 7 |
|  | Peripheral Blood Naïve B Cell | Immune | 4.12 | 0.07 | 7 |
|  | Renal Proximal Tubule Epithelial Cell | Primary Epithelial | 4.07 | 0.06 | 7 |
| Bladder | Fetal Ureteric Bud Epithelial Cell | Fetal Epithelial | 3.48 | 0.07 | NA |
|  | Prostate Epithelial Cell (RWPE-2) | Immortalized Epithelial | 3.38 | 0.05 | NA |

Bold font indicates high-confidence ( $PIP \geq 0.5$ ) biological contexts. NAs indicate the context was not included in any credible set.
S-LDSC: Stratified Linkage Disequilibrium Scores Regression; CT-FM: Cell Type Fine-Mapping; PIP: posterior inclusion probability.

Beyond these top signals, CT-FM identified 11 candidate (0.05 ≤ PIPs < 0.5) contexts across six cancers (**Table 2**). These included mammary basal epithelial cells in overall and ER+ breast cancer, and T47D mammary luminal adenocarcinoma epithelial cell line in ER+ breast cancer. Prostate cancer analysis revealed prostate adenocarcinoma (PRAD) bulk tissue, while colorectal cancer analyses identified stromal contributions from connective tissue mesenchymal stem cells and various fibroblast populations. Renal cancer analysis implicated peripheral blood naïve B cells and renal proximal tubule epithelial cells. For bladder cancer, fetal ureteric bud epithelial cells and RWPE-2 prostate epithelial non-malignant cell line emerged as candidate contexts, despite bladder cancer lacking significant S-LDSC associations.

A single 95% credible set was identified for each of these cancers except for bladder cancer, for which no credible set was formed given no combination of annotations reached the required 95% posterior probability threshold.

Detailed S-LDSC and CT-FM results for all annotations satisfying the S-LDSC q-value ≤ 0.05 or the CT-FM PIP ≥ 0.05 criterion are provided in **Table S2**.

### Fine-mapping biological contexts at single-variant resolution

A total of 837 high-confidence (PIPs ≥ 0.5) causal SNPs were identified by functionally informed SNP fine-mapping across all cancer types except esophageal cancer. Among these, 487 SNPs overlapping at least two annotations with positive enrichment z-scores were further tested (counts by cancer in **Table 3**).

**Table 3.** Summary of high-confidence (PIP ≥ 0.5) biological contexts identified by variant-level context fine-mapping.

| Cancer type | No. causal SNPs | No. SNPs tested | No. SNPs with high-conf contexts | No. high-conf contexts | Most frequent high-conf context | No. SNPs for most frequent context | No. quadruplets |
| --- | --- | --- | --- | --- | --- | --- | --- |
| Breast | 164 | 98 | 65 | 53 | Mammary Luminal Epithelial Cell | 22 | 70 |
| ER+ breast | 118 | 66 | 47 | 46 | Mammary Luminal Epithelial Cell | 20 | 60 |
| ER- breast | 32 | 22 | 2 | 16 | Breast Invasive Carcinoma (BRCA) | 2 | 7 |
| Endometrial | 22 | 13 | 3 | 3 | * | * | 0 |
| Ovarian | 10 | 9 | 0 | 0 | NA | NA | 0 |
| Prostate | 355 | 210 | 169 | 81 | Prostate adenocarcinoma (PRAD) | 86 | 283 |
| Colorectal | 15 | 5 | 3 | 3 | Colon Adenocarcinoma (COAD) | 2 | 1 |
| Esophageal | 0 | 0 | 0 | 0 | NA | NA | 0 |
| Lung | 18 | 8 | 1 | 13 | * | * | 13 |
| Renal | 88 | 46 | 29 | 43 | Kidney Renal Clear Cell Carcinoma (KIRC) | 17 | 47 |
| Bladder | 15 | 10 | 4 | 9 | Fetal Ureteric Bud Epithelial Cell | 2 | 8 |

Causal SNPs refer to high-confidence ( $PIP \geq 0.5$ ) causal SNPs detected by PolyFun+SuSiE SNP fine-mapping. Among these SNPs, those with at least two overlapping annotations were tested in CT-FM-SNP analysis. High-conf contexts refer to high-confidence ( $PIP \geq 0.5$ ) biological contexts detected by CT-FM-SNP. NAs indicate that CT-FM-SNP did not detect any high-confidence contexts for the cancer. Asterisks indicate that no single context can be considered most frequently detected (i.e., all contexts were detected in the CT-FM-SNP analysis of one SNP).

We performed biological context fine-mapping by focusing on each of these 487 SNPs and restricting analyses to their overlapping annotations through CT-FM-SNP. This variant-level approach yielded altogether 639 high-confidence (PIPs ≥ 0.5) SNP-context-cancer triplets across nine of the 11 cancer types analyzed, excluding ovarian and esophageal cancers (full list in **Table S3**). Prostate cancer yielded the greatest number of high-confidence associations (322 triplets involving 81 unique biological contexts), followed by overall breast cancer (119 triplets, 53 contexts), ER+ breast cancer (92 triplets, 46 contexts), and renal cancer (59 triplets, 43 contexts).

The variant-level signals were largely consistent with the genome-wide findings while capturing additional relevant high-confidence contexts (**Table 3** and **Table S3**). For both overall and ER+ breast cancer, mammary luminal epithelial cells represented the most frequently detected context, achieving high confidence in the analysis of 22 and 20 SNPs, respectively. ER– breast cancer predominantly implicated breast invasive carcinoma (BRCA) bulk tissue, although this association was observed for only 2 SNPs. Prostate cancer identified PRAD as the most frequent context across 86 SNPs, followed by VCaP across 85 SNPs. Colorectal cancer implicated COAD for 2 SNPs, while renal cancer most frequently identified KIRC across 17 SNPs. Bladder cancer demonstrated fetal ureteric bud epithelial cells as the most frequent context, detected for 2 SNPs. Endometrial and lung cancer yielded three and 13 high-confidence contexts, respectively—including COAD for endometrial and the MM.1S multiple myeloma cell line for lung—with each context detected for only one SNP.

Among 53 SNPs overlapping only one annotation with positive z-score, none were linked to high-confidence contexts detected in the genome-wide CT-FM analysis, though three were linked to candidate contexts (rs61752561, rs61735792, and rs7591218; all for prostate cancer and overlapping PRAD).

### {Regulatory SNP-context-gene-cancer} quadruplets

By integrating the SNP-context-cancer triplets with high-confidence SNP-gene pairs derived from multiple S2G linking strategies, we constructed a comprehensive set of 489 high-confidence {regulatory SNP-context-gene-cancer} quadruplets across nine cancers (counts by cancer in **Table 3**). **Table 4** highlights illustrative quadruplets for each cancer ranked by CT-FM-SNP PIP values, displaying up to five quadruplets per cancer where available. When multiple quadruplets existed for the same SNP-gene pair across different biological contexts, we present the context with the highest PIP value.

**Table 4.**
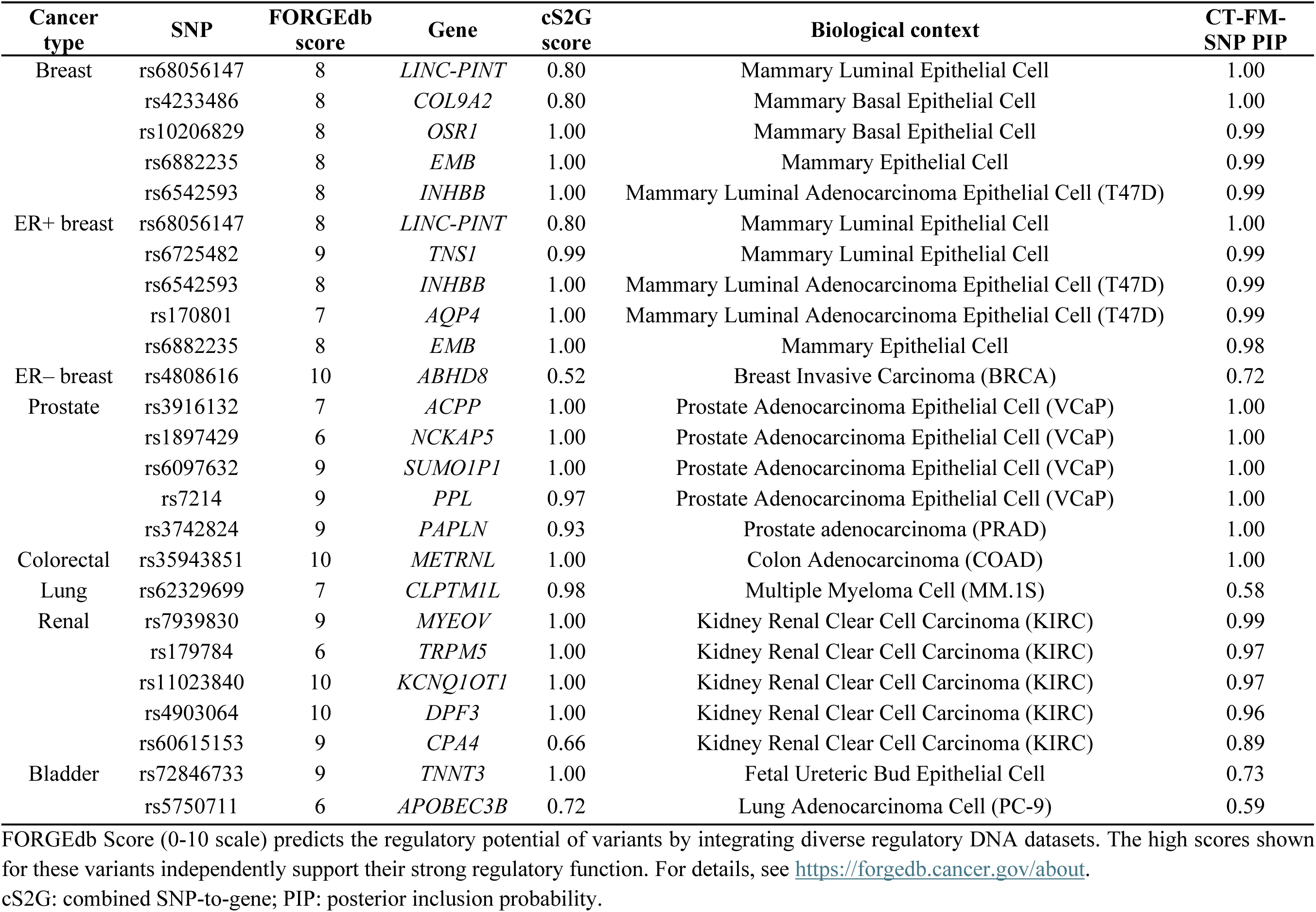
Examples of high-confidence regulatory quadruplets linking cancer risk variants to putative target genes and biological contexts.

This analysis yielded high-confidence regulatory mechanisms for particular SNPs within tissue-of-origin contexts for overall and ER+ breast cancer (e.g., rs68056147-*LINC-PINT*-mammary luminal epithelial cells), prostate cancer (e.g., rs3916132-*ACPP*-VCaP), colorectal cancer (only one quadruplet found: rs35943851-*METRNL*-COAD), and renal cancer (e.g., rs11023840-*KCNQ1OT1*-KIRC). For other cancers, the links between genes and contexts were less specific due to the absence of any single dominant signal—for example, bladder cancer SNP rs5750711 and its target gene *APOBEC3B* showed competing signals among seven contexts including PC-9 lung adenocarcinoma cell line and bladder urothelial carcinoma (BLCA) bulk tumor tissue, with CT-FM-SNP PIPs ranging from 0.51 to 0.59. Details of all identified quadruplets can be found in **Table S4**.

### Sensitivity analyses excluding bulk tumor annotations

When excluding TCGA bulk tumor annotations, CT-FM consistently identified mammary luminal epithelial cells as the high-confidence biological context for both overall breast cancer and its ER+ subtype (PIP = 0.62-0.75), and VCaP as the high-confidence context for prostate cancer (PIP = 1.00). No high-confidence contexts were identified for the remaining cancer types (**Table S5**). In the corresponding CT-FM-SNP analysis, the most frequently detected contexts for most cancer types remained concordant with the primary analysis, except for renal cancer, where the predominant context shifted from KIRC to renal proximal tubule epithelial cells and peripheral blood naïve B cells (**Table S6**). Complete {regulatory SNP-context-gene-cancer} quadruplets derived from this analysis are provided in **Table S7**.

## Discussion

In this study, we employed an integrative statistical fine-mapping framework to systematically elucidate the cellular origins of genetic susceptibility across 11 solid cancers. By integrating large-scale GWAS data with an atlas of approximately 1,500 context-specific *cis*-regulatory elements, our approach advanced beyond traditional heritability partitioning to identify potential cancer-relevant biological contexts. A central contribution of our work lies in the substantial refinement of putative causal signals achieved through statistical fine-mapping. Whereas conventional enrichment analysis (i.e., S-LDSC) implicated 141 context-specific annotations, our fine-mapping process, which explicitly models annotation co-regulation, distilled these associations into four high-confidence biological contexts for breast, prostate, colorectal, and renal cancers, respectively. This represents an advancement in specificity, enabling the disentanglement of putative causal contexts from bystander signals attributed to shared regulatory programs.

Our genome-wide and variant-specific analyses consistently converged on findings that reinforce established biological paradigms in cancer etiology. At a broad level, genome-wide analyses identified tissue-of-origin epithelial contexts as the predominant drivers of heritable risk for these cancers. This pattern was exemplified by the prioritization of mammary luminal epithelial cells for both overall and ER+ breast cancer, consistent with their established role as the presumptive cells of origin for breast malignancies.^39,40^ The strong signal for the VCaP epithelial cell line, derived from a prostate cancer vertebral metastasis, suggests that the regulatory architecture disrupted by germline risk variants is not only critical for disease initiation but may also be retained and relevant in advanced, metastatic states.^41,42^ The prioritization of bulk tumor tissues for colorectal and renal cancers is consistent with this observation. The bulk tumor may act as a proxy for the true cell of origin, retaining the regulatory signatures shaped by germline variants through malignant transformation.^43^ The signal may also arise from the collective contribution of non-cancerous cell types—such as resident immune and stromal populations—that coinhabit tumors.^8^ Both interpretations point to the *in vivo* tissue environment as the critical context for understanding how germline variants confer cancer risk. Variant-level fine-mapping closely aligned with these results, with the same epithelial and bulk tumor contexts frequently emerging as the prioritized context for individual risk variants identified through functionally informed fine-mapping, while also providing additional insights at higher resolution. For instance, in prostate cancer, the bulk prostate tumor tissue—a candidate signal in the genome-wide analysis—emerged as the most frequent high-confidence context for individual SNPs alongside VCaP. Similarly, for ER– breast cancer, which lacked a genome-wide signal, the variant-level approach successfully validated bulk breast tumor tissue as a key etiologic context.^44^ These findings demonstrate the complementary value of multi-scale analytical approaches in dissecting cancer heritability.

Beyond the observations aligning with established cell– and tissue-of-origin biology, our analysis also uncovered less-obvious yet biologically plausible contexts warranting further investigation. Our results underscore the critical role of the TME;^8^ for instance, the implication of various stromal cell types in colorectal cancer, while not colonic in origin themselves, likely points to a shared, pan-fibroblastic regulatory program, or reflects these annotations acting as strong statistical proxies for the true causal cell populations within the colon’s stroma. The contribution of TME may also be reflected in the implication of immune cell types, such as naïve B cells, in renal cancer, corroborating the importance of the immune response in cancer susceptibility.^8^ Furthermore, our discoveries point to potential for cross-tissue etiologies, suggesting shared biological programs underlying cancer risk across different organs. Notable examples include a compelling set of signals for bladder cancer supported by both prostate epithelial cells and fetal ureteric bud cells, consistent with their common developmental origin and related epithelial lineages;^45^ and the association of bulk endometrial tumor tissue (UCEC) with ER+, but not ER–, breast cancer risk, corroborating known shared hormonal signaling pathways.^46^

To translate these biological contexts into more specific molecular mechanisms, we mapped our context-specific causal variants to their putative regulatory genes using a heritability-based framework combining various S2G linking strategies.^38^ A review of these connections reveals that many of the mapped genes have well-documented or plausible roles in cancer biology. Examples include the established tumor-suppressive function of *LINC-PINT* in breast cancer^47^ and oncogenic role of *CLPTM1L* in lung cancer,^48^ as well as emerging connections such as *DPF3*, a chromatin remodeling factor previously implicated in kidney cancer cell migration,^49^ and *TNS1*, whose role in cell invasiveness may contribute to breast cancer risk.^50^ Integrating these gene links with our prior results allowed us to construct 489 {regulatory SNP-context-gene-cancer} quadruplets. The credibility of these proposed mechanisms is underscored by cases where our framework points to interactions already supported by external evidence. For example, our quadruplet for prostate cancer links genetic risk to the *ACPP* gene within prostate epithelial cells— the very cells where prostatic acid phosphatase (ACPP) is known to be synthesized.^51,52^ Similarly, our link between *KCNQ1OT1* and genetic susceptibility to renal cancer in bulk kidney tumor tissue is supported by the gene’s known high expression in KIRC tumors.^53^ Beyond reinforcing known interactions, these quadruplets propose novel testable mechanisms, each defining a full regulatory path from variant to gene, context, and ultimately, to cancer. Interestingly, our results also illuminate potential mechanisms of pleiotropy, revealing how loci near *TERT*, *TET2*, *FGFR2*, *CDKN1A*, *RHOD*, and *ANXA9* are implicated across multiple malignancies. For example, the *RHOD* locus (11q13.2) is implicated in both ER+ breast cancer (mammary luminal epithelial cells) and prostate cancer (VCaP and PRAD) specifically through variant rs114895445, and the *CDKN1A* locus (6p21.2) is linked to both prostate (VCaP) and renal cancer (KIRC) through variant rs762624 (illustrated as an example in **Figure 3**), highlighting cancer-shared genetic pathways that may be activated through distinct biological contexts.

**Figure 3.**
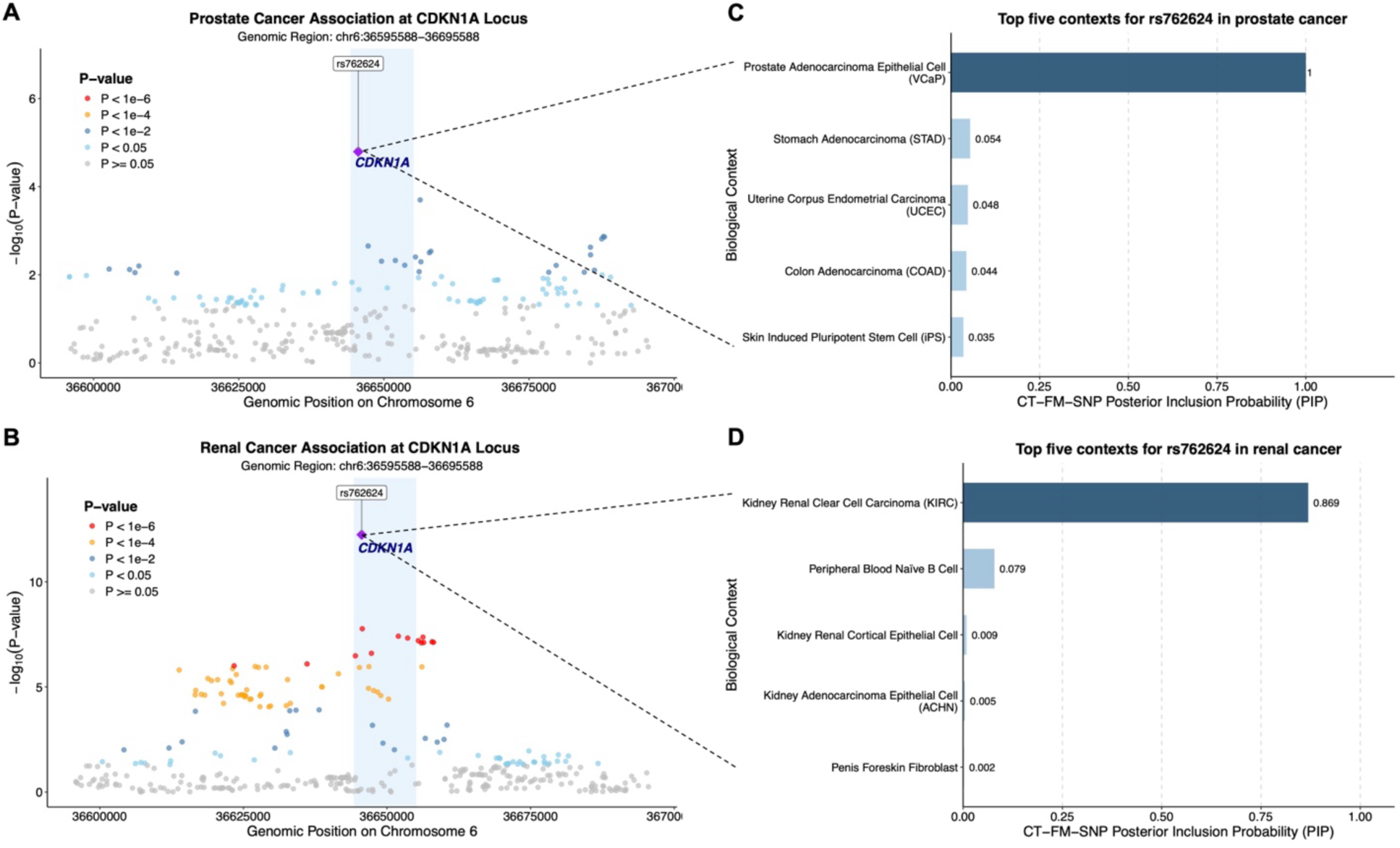
Context-specific pleiotropy at *CDKN1A* locus in prostate and renal cancer. (**A**, **B**) Regional association plots for prostate and renal cancer at *CDKN1A* locus (6p21.2), showing genetic variant associations within a 50 kb window surrounding the putatively causal variant rs762624 (purple diamond; PolyFun+SuSiE posterior inclusion probability [PIP] ≥ 0.5). Light blue shaded regions delineate *CDKN1A* gene boundaries (chr6:36,644,240-36,655,109, GRCh37/hg19; NCBI Gene). Y-axes show the *P*-value of association on a –log_10_ scale. (**C**, **D**) The top five biological contexts for rs762624 as prioritized by CT-FM-SNP analysis in prostate and renal cancer, respectively, and ranked by PIP.

Several limitations warrant consideration when interpreting our findings. Our analytical power is inherently dependent on the quality and scope of its inputs: specifically, the statistical strength of the underlying GWAS, the external LD reference panel, and the comprehensiveness of our functional annotation library. The dependency on well-powered GWAS likely explains the limited results for cancers with smaller sample sizes, such as esophageal cancer (n=4,112 cases), and consequently constrained our study’s scope to these 11 solid cancers, precluding analysis of other malignancies for which large-scale summary statistics are not yet available (or publicly accessible). We were also limited by the lack of availability of large-scale GWAS from non-European ancestry populations. Future investigations can apply this framework in a multi-ancestry setting, harnessing shared genetic effects and diverse LD patterns across populations to enhance statistical power and fine-mapping resolution.^54^ Furthermore, if the truly causal cell type or precise biological context is not included in our reference panel, our context fine-mapping model may identify the best available proxy cell types or tissues. Therefore, an identified context should be interpreted as representing a broader relevant biological program rather than a literal, definitive cellular source. This same principle applies to the fine-mapped variants themselves, as the true causal SNP may be absent from our analysis due to initial SNP reference panel limitations or filtering. Consequently, all findings presented here are data-driven hypotheses that necessitate at least two crucial avenues of follow-up: first, replication and expansion in larger, more diverse GWAS cohorts and annotation datasets —including single-cell data from relevant tumor types to deconvolve bulk tissue signals—and second, extensive laboratory functional validation to confirm their biological mechanisms before any etiological relevance can be considered.

## Conclusion

To conclude, we applied an integrative statistical fine-mapping framework that leverages *cis*-regulatory element annotations to prioritize context-specific genetic risk mechanisms across 11 solid cancers. By progressing systematically from genome-wide patterns to single-variant effects, our analysis disentangled putative causal contexts from correlated signals, revealing both established and novel regulatory mechanisms. These findings advance our understanding of the genetic susceptibility to different cancers and provide a potential roadmap to guide experimental studies aimed at translating genetic discoveries into mechanistic and, ultimately, clinical insights.

## Acknowledgments

We gratefully acknowledge all researchers and consortia who have made GWAS summary statistics and regulatory annotations publicly available or shared data through collaborative arrangements. The willingness of the scientific community to share these valuable datasets and annotations is essential for advancing our understanding of complex trait genetics and regulatory mechanisms, and enables secondary analyses such as this study. We thank the original study participants and investigators whose contributions made these datasets possible.

## Data availability

All publicly available GWAS summary statistics following standardized quality control procedures have been uploaded to https://zenodo.org/records/15676245, except for the bladder cancer dataset, which has controlled access through dbGAP phs003342.v1.p1 for research related to bladder cancer. Processing procedures and detailed data characteristics are also documented at https://github.com/xueyaowunci/cancer-context-fine-mapping. Analysis scripts and source of all cis-regulatory element annotations can be found at https://github.com/ArtemKimUSC/CTFM.

## Authors’ contributions

XW and PK designed the study. XW collected and processed all GWAS summary statistics with the help of SL. AK curated the CT-FM software and provided all context-specific annotation data. XW performed the primary analyses with the assistance of AK, PK, and NM. XW, PK, CEB, SL, AK, NM, TOM, DR, TD, SK, NR, and LPO interpreted the results. XW drafted the manuscript with significant input from PK. All co-authors contributed to review and revision of the manuscript.

## Ethics declaration

This study utilized exclusively summary-level GWAS data from previous studies. No individual-level genetic or phenotypic data were accessed or analyzed. As all data used were aggregate summary statistics that do not contain identifiable information about individual participants, this research did not require institutional review board approval or ethics committee oversight. The original studies from which the summary statistics were derived had obtained appropriate ethical approvals and participant consent as reported in their respective publications.

