## Supplementary Methods for "Prioritizing context-specific genetic risk mechanisms in 11 solid cancers"

**Supplementary Methods: Curation of context-specific SNP-annotations**

Our dataset of context-specific regulatory SNP annotations comprised 1,473 total annotations, including 927 SNP annotations previously curated for the CT-FM framework and 546 additional annotations specifically curated to enhance CT-FM’s applicability to cancer trait analysis. The annotations were derived from five primary sources: the fourth phase of ENCyclopedia of DNA Elements (ENCODE4) (N=842 annotations), Cis-element Atlas (CATlas) (N=222), Activity-by-Contact (ABC) model (N=104), Epigenome Integration across Multiple Annotation Projects (Epimap) (N=282), and The Cancer Genome Atlas (TCGA) (N=23).

For ENCODE4, we used cell-type-specific *cis*-regulatory elements defined by a combination of experimental assays, including DNase-seq for chromatin accessibility and ChIP-seq for histone modifications. These annotations were manually curated by excluding tissue-level data to maintain cell-type specificity. For each resulting *cis*-regulatory element file, we excluded regulatory marks corresponding to “low DNase” and “Unclassified” states. ENCODE4 genomic coordinates were converted from hg38 to hg19 using the UCSC liftOver tool. CATlas data, a comprehensive atlas of chromatin accessibility peaks derived from ATAC-seq, was retrieved and converted to hg19 using liftOver without additional filtering. For ABC, we retrieved gene-enhancer prediction datasets. The ABC model is a computational framework that predicts enhancer-gene links by integrating measures of enhancer activity (e.g., H3K27ac ChIP-seq) with 3D chromatin contact data (e.g., Hi-C). We excluded tissue-level ABC predictions to maintain cell-type resolution. For EpiMap, we focused on 282 enhancer-gene regulatory maps corresponding to cancer-related and immune-related cell lines; these maps are primarily defined by ChIP-seq data for various histone modifications that denote enhancer activity. For TCGA, cancer type-specific chromatin accessibility peak calls were converted to hg19 using liftOver and used without additional filtering.

All resulting BED files were sorted using the sortBed function of bedtools and validated for formatting errors using the BEDOPS --ec --merge function. The final dataset contained 1,473 context-specific *cis*-regulatory SNP annotations with an average genome coverage of 31.6 Mb per annotation. Detailed annotation names, genomic coverage, and source information for each annotation are provided in **Table S1**.
